# Gene Expression Analysis of Patients with Prostate Cancer vs Benign Prostate Hyperplasia

**DOI:** 10.1101/2022.12.21.22283808

**Authors:** Mahdi Saeedi

**Affiliations:** University of North Dakota

**Keywords:** Benign Prostate Hyperplasia, Differential Expression, Gene Expression, Prostate Cancer

## Abstract

Prostate cancer is associated with nearly 4% of all cancer-related deaths in men. To better understand the types of genes that are highly expressed in patients with prostate cancer and benign prostate hyperplasia, this study analyzes the raw gene expressions of 36 patients. And compares expressions of prostate epithelial stem cells vs. prostate epithelial transit amplifying cells. 1347 differentially expressed genes were found in prostate epithelial stem cells, and 71 differentially expressed genes were found in prostate epithelial transit amplifying cells. Genes USP10 and SOCS3 were found to be differentially expressed and are found to be associated with prostate cancer.

## I. Introduction

Gene expression analysis is a technique used to study the activity of genes in a cell or tissue. It involves measuring the levels of RNA or proteins produced by a gene, as well as other factors that can affect gene expression. This information can be used to understand how genes are regulated, how they interact with each other, and how they contribute to the development and function of an organism. By analyzing gene expression patterns, researchers can gain insight into the underlying mechanisms of biological processes and disease. This knowledge can be used to develop new therapies and treatments, as well as to improve our understanding of biology and medicine. In this study, we focus on the gene expression of patients with prostate cancer and benign prostate hyperplasia. The specific study we use is “Transcription Profiling by array of human prostate cancer stem cells” from the University of York by Richard Birnie [1].

Transcription profiling by array is a laboratory technique that is used to study gene expression in cells. It involves using DNA microarrays, which are small glass or silicon slides that are coated with DNA probes that correspond to specific genes. In the case of human prostate cancer stem cells, transcription profiling by array would involve isolating a population of prostate cancer stem cells from a patient with prostate cancer and then using DNA microarrays to study the expression of specific genes in these cells. This technique can be useful for identifying which genes are active or “expressed” in prostate cancer stem cells, as well as for comparing gene expression patterns between different samples. It can provide insights into the mechanisms that drive the development and progression of prostate cancer, which may ultimately lead to the development of new treatment strategies.

Prostate cancer is a type of cancer that shows up in the prostate. The prostate is part of the male reproductive system located below the bladder and in front of the rectum. The prostate is about a size of a mature walnut. The prostate is responsible for the portion of the fluid that makes up the semen. As men age, the prostate tends to increase in size, which can lead to a host of issues. The issues range from decreased urine flow, blood in urine, blood in semen, bone pain, unintentional weight loss, and erectile dysfunction. The condition where the prostate grows in size is referred to as benign hyperplasia [2]. Not all changes in the prostate are cancers. Sometimes inflammation can be caused by bacterial infections [3].

Our raw dataset contains 36 male patients. The gene expressions are extracted from prostate epithelial stem cells and prostate epithelial transit amplifying cells. This study examines the gene expressions in the two mentioned cells to identify differentially expressed genes in patients with prostate adenocarcinoma vs. benign prostatic hyperplasia.

## II. Disease Background

Prostate cancer is a type of cancer that develops in the prostate, a small gland in the male reproductive system. The prostate gland is located just below the bladder and in front of the rectum. It is responsible for producing fluid that helps to nourish and transport sperm during ejaculation.

Prostate cancer typically affects older men, i.e., older than 65 years of age. Nearly 4% of all deaths caused by cancer for men are prostate cancer. [4]. It is one of the most common types of cancer in men, and the risk of developing it increases with age.

Symptoms of prostate cancer may include difficulty urinating, frequent urination, and pain or burning during urination. However, these symptoms can also be caused by other conditions, such as an enlarged prostate or a urinary tract infection, so it is important to see a doctor for a proper diagnosis.

Prostate cancer is typically diagnosed through a combination of a physical exam, a blood test called a prostate-specific antigen (PSA) test, and a biopsy. Treatment options may include surgery, radiation therapy, and hormone therapy, among others. The choice of treatment will depend on the stage and grade of cancer, as well as the overall health of the patient.

It is important to catch prostate cancer early, as it is typically more treatable in its early stages. Regular checkups with a doctor, including a prostate exam and PSA test, can help to detect prostate cancer early.

It is generally believed that prostate cancer develops as a result of a combination of genetic and environmental factors, including age, diet, and exposure to certain chemicals. Many prostate cancers are detected on the basis of elevated plasmatic levels of prostate-specific antigen (PSA > 4 ng/mL), which is a glycoprotein expressed in prostate tissue [4]. Research has identified certain genetic mutations that may increase the risk of developing prostate cancer, but these mutations are not present in all cases of the disease. In addition, research has also identified certain dietary and lifestyle factors that may increase the risk of prostate cancer, such as a diet high in red meat and processed foods and a sedentary lifestyle. It is important to note that while certain factors may increase the risk of developing prostate cancer, they do not necessarily cause the disease. More research is needed to fully understand the underlying causes of prostate cancer.

There are two specific conditions we are focused on in this study. The first one is Benign prostatic hyperplasia (BPH) is a non-cancerous condition that affects the prostate gland. In BPH, the prostate gland becomes enlarged, which can cause a number of urinary symptoms, such as difficulty urinating, frequent urination, and a weak or interrupted urine flow. These symptoms may be mild at first, but they can worsen over time and may lead to complications, such as urinary tract infections or bladder damage.

BPH is a common condition that typically affects older men. It is estimated that roughly more than 50% of men aged over 60 years of age suffer from BPH [5]. It is not the same as prostate cancer, which is a separate condition that develops in the prostate gland. However, it is important to see a doctor for a proper diagnosis and treatment, as BPH can sometimes be confused with prostate cancer.

BPH is typically treated with medication, such as alpha-blockers [6], which can help to relax the muscles in the prostate and make it easier to urinate. In some cases, surgery may be necessary to remove part of the prostate gland. It is important to work with a doctor to determine the best course of treatment for BPH.

Prostate adenocarcinoma is a type of cancer that develops in the prostate gland. Prostate adenocarcinoma is the most common type of prostate cancer, accounting for about 95% of all cases. It is slow-growing cancer that typically affects older men and is rare in men under the age of 40 [7].

Symptoms of prostate adenocarcinoma may include difficulty urinating, frequent urination, and pain or burning during urination. However, these symptoms can also be caused by other conditions, such as an enlarged prostate or a urinary tract infection, so it is important to see a doctor for a proper diagnosis [8].

Prostate adenocarcinoma is typically diagnosed through a combination of a physical exam, a blood test called a prostate-specific antigen (PSA) test, and a biopsy. Treatment options may include surgery, radiation therapy, and hormone therapy, among others. The choice of treatment will depend on the stage and grade of cancer, as well as the overall health of the patient [9]. It is important to catch prostate adenocarcinoma early, as it is typically more treatable in its early stages. Regular checkups with a doctor, including a prostate exam and PSA test, can help to detect prostate cancer early.

The next important topic we need to cover is Stem cells. They are cells that can self-renew and give rise to other cell types. They are undifferentiated cells, meaning they have not yet specialized into specific cell types, such as nerve cells, muscle cells, or blood cells [10].

Stem cells are important because they have the potential to develop into many different cell types in the body. This ability to give rise to other cell types makes them unique and allows them to play a key role in the repair and regeneration of tissues and organs.

There are two main types of stem cells: embryonic stem cells and adult stem cells. Embryonic stem cells are derived from early-stage embryos and have the ability to give rise to any cell type in the body. Adult stem cells, on the other hand, are found in various tissues and organs in the body and have a more limited ability to give rise to different cell types. Stem cells are a topic of active research and have the potential to be used in the development of new therapies for a variety of diseases.

In this study, we focus on stem cells that contain CD133 and lack it. CD133 is a protein that is expressed on the surface of certain cells, including cancer cells. It is often used as a marker for cancer stem cells, which are a subpopulation of cancer cells that have the ability to self-renew and give rise to other cancer cells. In some types of cancer, such as certain types of brain and blood cancers, CD133+ cancer cells have been shown to be resistant to chemotherapy and radiation therapy, making them a potential target for new treatment strategies [11].

Cells that lack CD133 are known as CD133-stem cells. In our study patients with that are marked as CD133-contain Alpha2integrin, which is a type of protein that is found on the surface of certain cells, including cancer cells. It is a member of a larger family of proteins called integrins, which play a role in cell adhesion and migration.

In cancer cells, alpha2integrin has been shown to be involved in the process of angiogenesis, which is the formation of new blood vessels [12]. This is important because cancer cells require a steady supply of nutrients and oxygen from the bloodstream to grow and spread.

Low levels of alpha2integrin on the surface of cancer cells have been associated with a more aggressive form of cancer and a worse prognosis for the patient. This is because low levels of alpha2integrin may allow cancer cells to more easily detach from the original tumor and migrate to other parts of the body, where they can form new tumors. More research is needed to fully understand the role of alpha2integrin in cancer and how it may be targeted as part of a treatment strategy.

It is likely that gene expression profiles in CD133+ and alpha2integrin low cell populations from patients with prostate cancer would differ from those of patients with benign prostatic hyperplasia (BPH), as these are two distinct conditions that affect the prostate gland.

Two types of cells are extracted from patients, Prostate Epithelial Transit amplifying cells and prostate epithelial stem cells. Prostate epithelial transit amplifying cells are a type of cell that is found in the prostate gland. They are called “transit amplifying” cells because they have the ability to divide and give rise to other cells, including both stem cells and more differentiated cells [13]. Prostate epithelial transit amplifying cells are important for maintaining the health and function of the prostate gland. However, they have also been implicated in the development of prostate cancer. In some cases, these cells may undergo changes that allow them to become cancerous and give rise to prostate cancer cells.

Prostate epithelial stem cells are a type of adult stem cell that is found in the prostate gland. They are responsible for maintaining the health and function of the prostate gland by replacing cells that are lost through normal wear and tear or injury.

Prostate epithelial stem cells are important for maintaining the health and function of the prostate gland. However, they have also been implicated in the development of prostate cancer [14]. In some cases, these cells may undergo changes that allow them to become cancerous and give rise to prostate cancer cells.

## III. Statistical Analysis

In this work, we are using the “maEndToEnd” package [15] to extract and analyze raw gene expressions. The accession code for our particular study is E-MEXP-993 and can be accessed via ArrayExpress [1]. We first examine some basic statistics about our raw dataset. Next, we will jump into extracting useful features and performing statistical analysis on gene expressions.

Our raw dataset contains 37 columns. For our purposes, we extract the following four columns. They are Source Name, Disease State, Individual Expression value, and Differentiation State. The 36 male patients in our study are either dealing with Prostate Adenocarcinoma or Benign Prostatic Hyperplasia, the differentiation states of which are prostate epithelial stem cells and prostate epithelial transit amplifying cells. Table 1 depicts a subsample of our raw dataset.

**Table 1.**
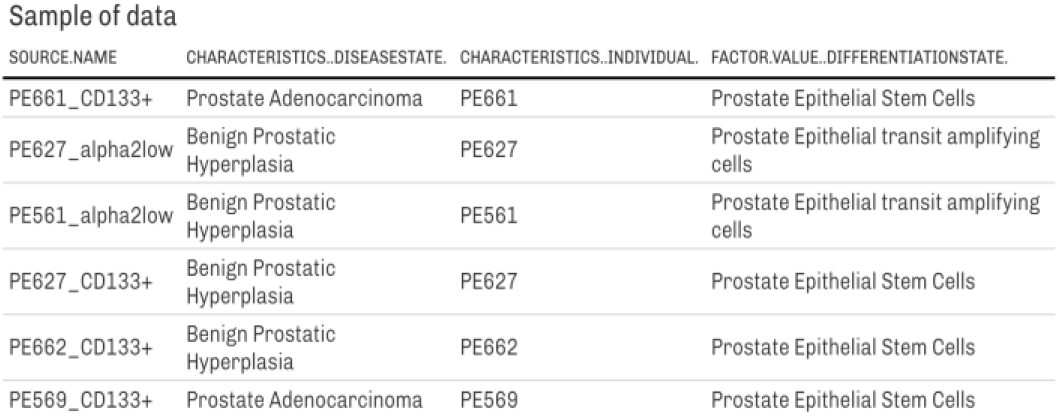
Sample of Data

Next step, we examine the intensity values of each cell. In table 2, we see a wide range of values in for our raw expression, which tells us that we need to normalize the values in order to perform statistical analysis. We normalize the values on a log2 base which enables us to better compare the values [16].

**Table 2.** Raw Intensity Values

| PE661CD133.CEL | PE627Alpha2Low.CEL | PE561Alpha2Low.CEL | PE627CD133.CEL |
| --- | --- | --- | --- |
| 147 | 96 | 108 | 92 |
| 12743 | 5808 | 8845 | 7102 |
| 199 | 74 | 103 | 80 |
| 13293 | 6464 | 9082 | 7344 |

We have to normalize this data by applying log transformation, which helps us perform linear analysis of our data. Table 3 shows a sample of the log2 normalized data.

**Table 3.** Normalized Expression Values

| PE661CD133.CEL | PE627Alpha2Low.CEL | PE561Alpha2Low.CEL | PE627CD133.CEL |
| --- | --- | --- | --- |
| 7.199672 | 6.584963 | 6.754888 | 6.523562 |
| 13.637417 | 12.503826 | 13.110646 | 12.794010 |
| 7.636625 | 6.209453 | 6.686501 | 6.321928 |
| 13.698379 | 12.658211 | 13.148794 | 12.842350 |

This enables us to apply principle component analysis on our raw data to extract relationships between the Disease state and differentiation state. Examining the first PCA plot in figure 3 of the raw log2 scaled data. We cannot differentiate between the disease or the disease State based on the current values at hand. We have to further analyze and normalize our data to identify outliers to be able to develop a useful visualization.

**Figure 1.**
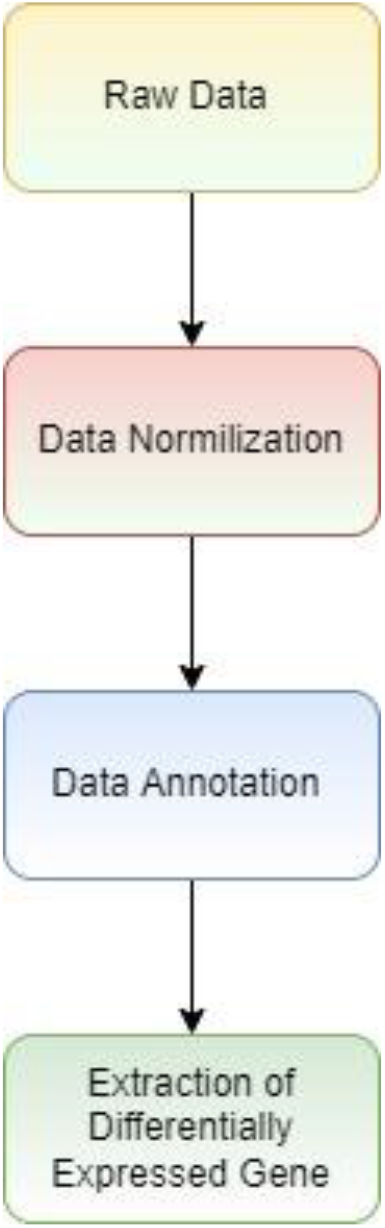
study flow

**Figure 2.**
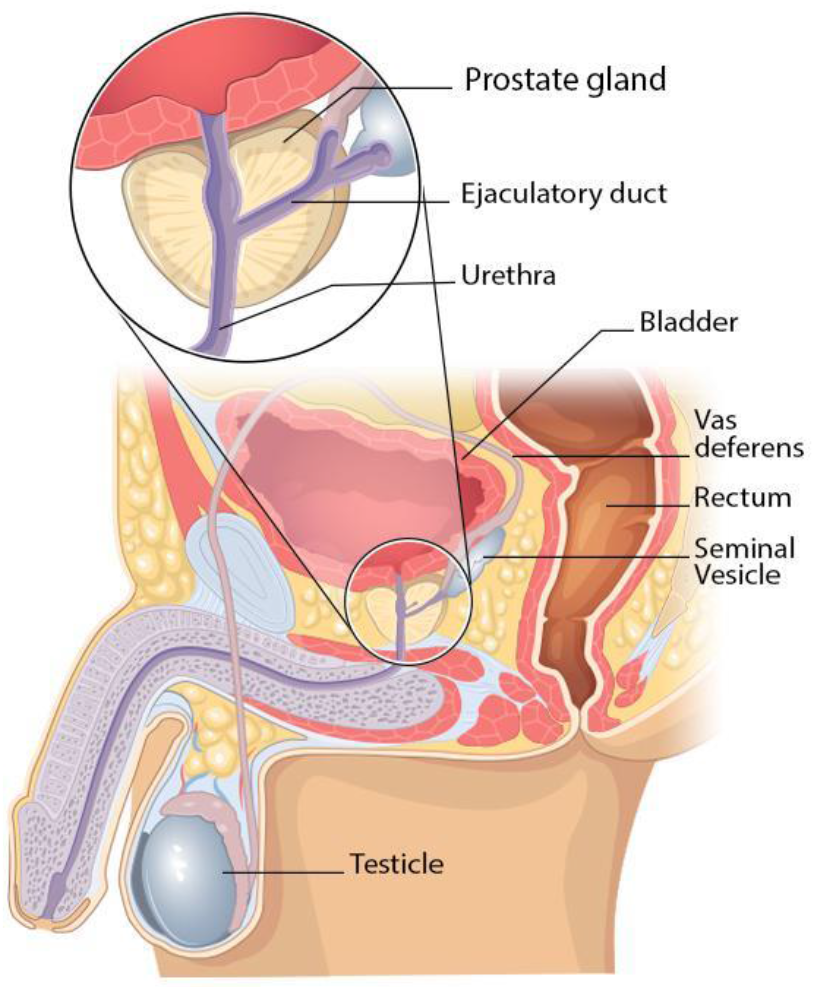
Male Genitalia Diagram [24]

**Figure 3.**
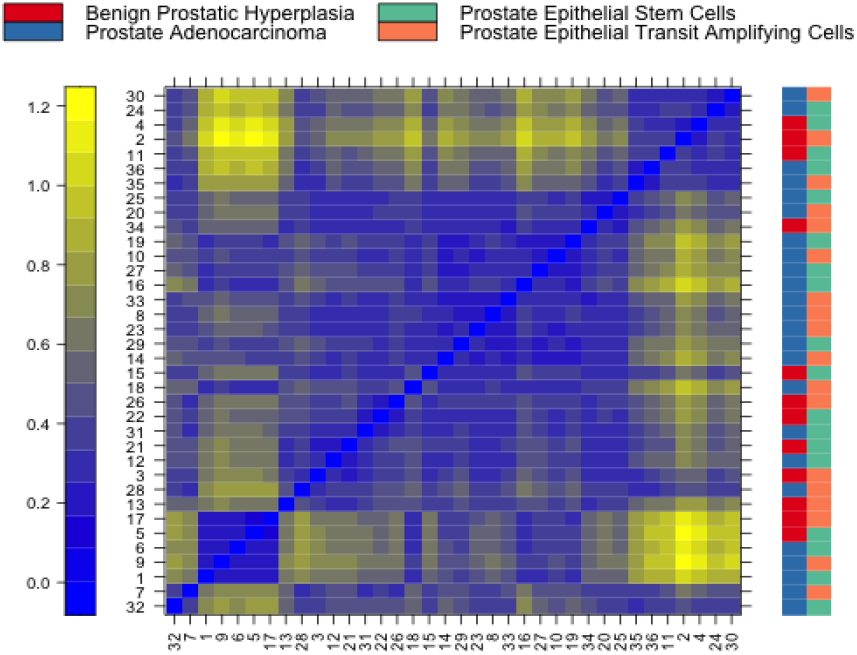
Heatmap of distances between arrays

Given that our first PCA results are not helpful, we can examine the distance between the arrays in order to look for outliers. Figure 4 is a false color heatmap of the distances between arrays. The color scale is chosen to cover the range of distances encountered in the dataset. Patterns in this plot can indicate clustering of the arrays either because of intended biological or unintended experimental factors (batch effects). The distance d_ab_ between two arrays, a, and b, is computed as the mean absolute difference (L1-distance) between the data of the arrays (using the data from all probes without filtering). In the formula, dab = mean | M_ai_ - M_bi_ |, where Mai is the value of the i-th probe on the a-th array. Outlier detection was performed by looking for arrays for which the sum of the distances to all other arrays, S_a_ = Σb d_ab_ was exceptionally large. No such arrays were detected.

**Figure 4.**
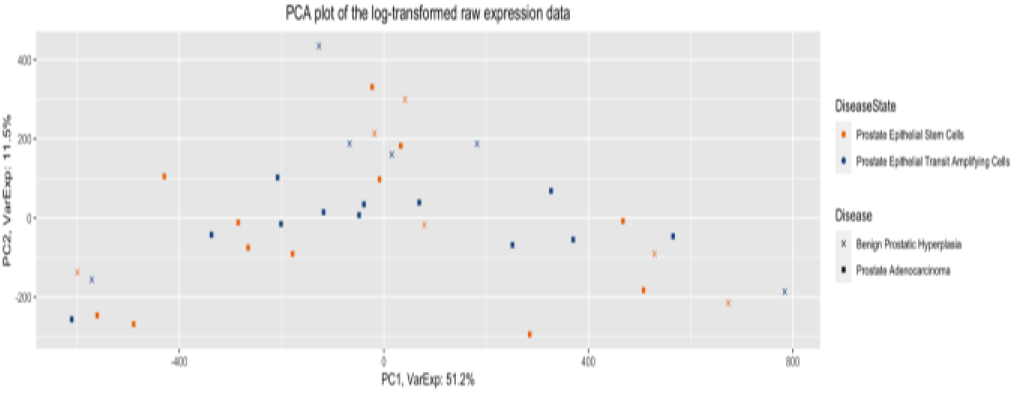
PCA plot Log-Transformed Raw Expression

One of the useful methods of analyzing expression values is boxplots. We can further analyze the range of expressions in our raw data using a boxplot. From this plot, we can see an acceptable range of values in our raw expression data. However, given the fact that our first PCA was not useful in yielding hints to differentiate between disease and disease state, we continue to normalize the data. Figure 5 shows the intensity of raw data. From the range of the bars, we can see that all the boxes are in comparable ranges.

**Figure 5.**
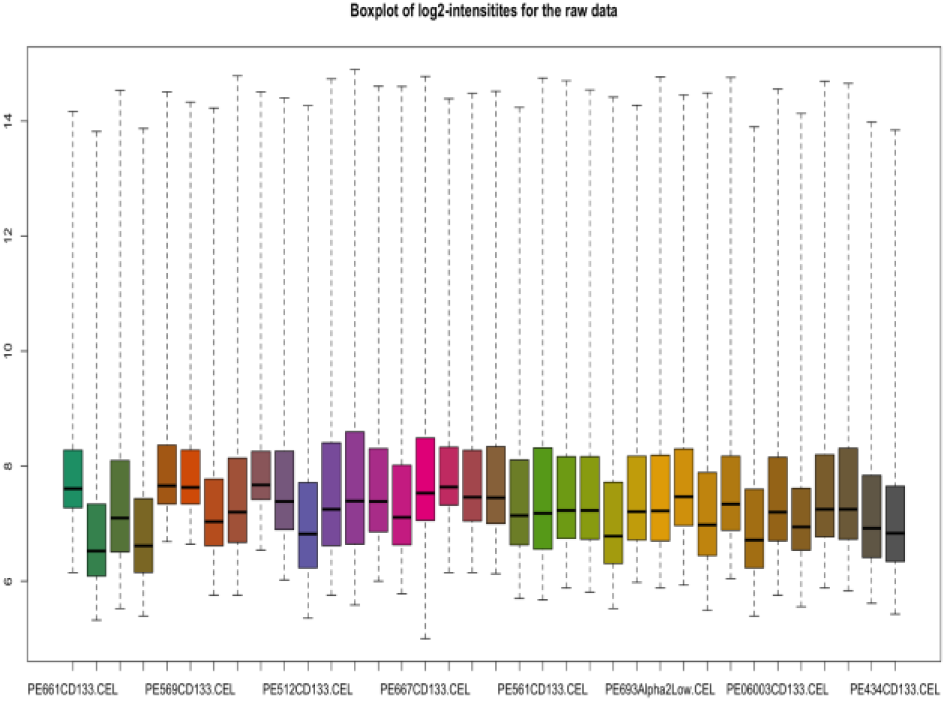
log2 intensities of raw data

Due to numerous obscure sources of variance, normalization across arrays is required in order to compare results from various array hybridizations. Reverse transcription efficiency issues, labeling or hybridization issues, physical issues with the arrays, reagent batch effects, and laboratory circumstances are a few of these. The background-corrected and normalized intensities of all probes for each gene must be combined into a single value that approximates a quantity proportionate to the amount of RNA transcript.

Figure 6 shows a density plot of the standard deviation of the intensities across arrays on the *y*-axis versus the rank of their mean on the *x*-axis. The red dots, connected by lines, show the running median of the standard deviation. After normalization and transformation to a logarithm(-like) scale, one typically expects the red line to be approximately horizontal, that is, show no substantial trend. In this case, a hump on the right hand of the x-axis can be observed and is symptomatic of saturation of the intensities.

**Figure 6.**
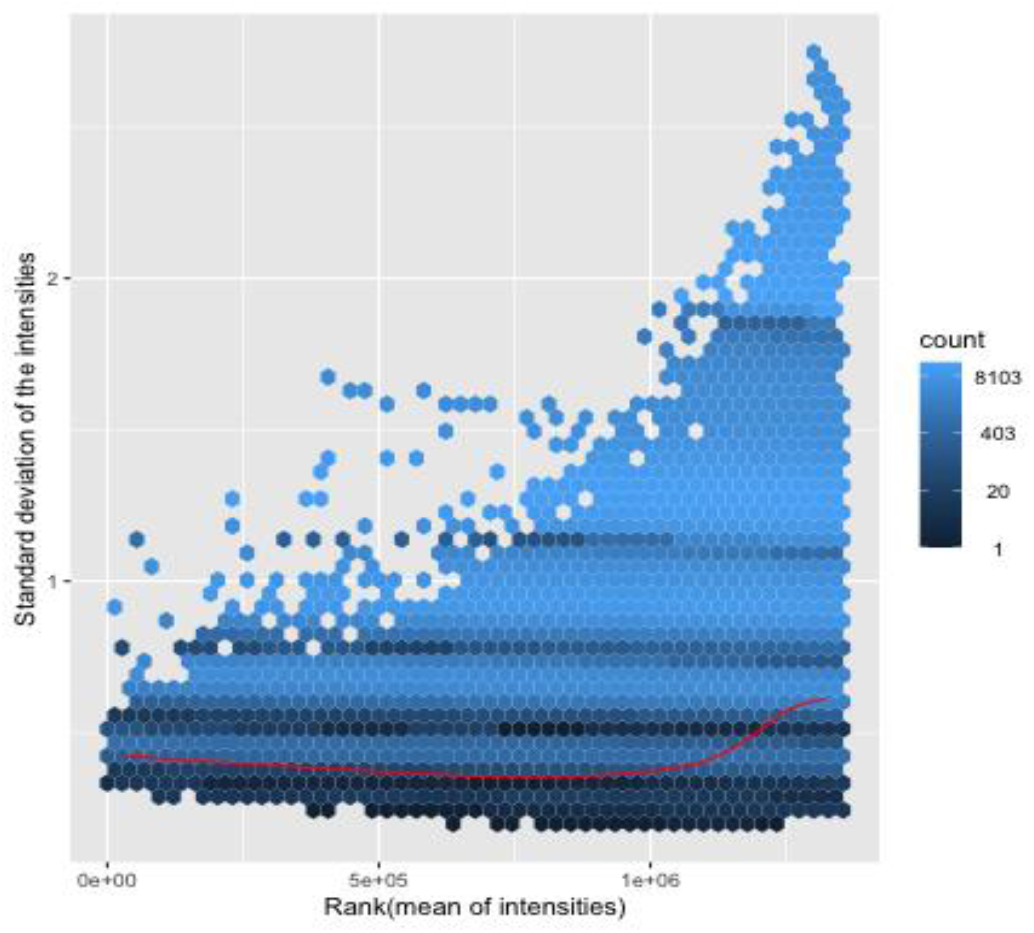
std of deviations

Background adjustment comes after the initial import and quality evaluation in the processing of microarray data. This is crucial because non-specific hybridization and optical detection system noise accounts for a percentage of the reported probe intensities. To provide precise measures of particular hybridization, measured intensities must be corrected.

We use the RMA algorithm without prior normalization [17]. We then calculate the relative log expression by calculating the median log2 intensity [18] of every transcript across all arrays. After performing the mentioned calculations, we can obtain a boxplot of log2 expression deviations. RMA is usually a good default choice. RMA shares information across arrays and uses the versatile quantile normalization method that will make the array intensity distributions match. Figure 7 y-axis displays for each microarray the deviation of expression intensity from the median expression of the respective single transcripts across arrays. Boxes with larger extensions indicate an unusually high deviation from the median in transcripts, suggesting a difference from other arrays. Boxes with shifts in y-direction indicate a systematically higher or lower expression of the majority of transcripts in comparison to most of the other arrays. The cause of this could stem from quality issues or batch effects. In this boxplot, we can potentially flag PE512Alpha2Low as an outlier.

**Figure 7.**
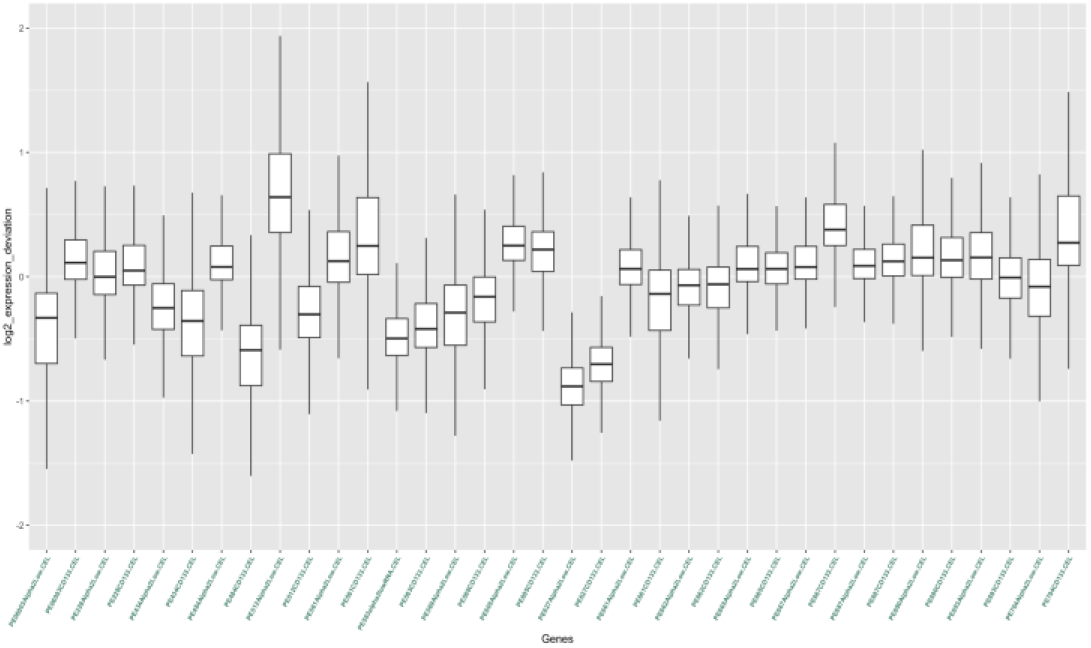
log2 expression deviations

After normalizing our raw expression data, we perform principle component analysis again. From Figure 8, we see that the first principle component can distinguish between the disease state. Indicating differential expression between disease states can be a dominant source of variation.

**Figure 8.**
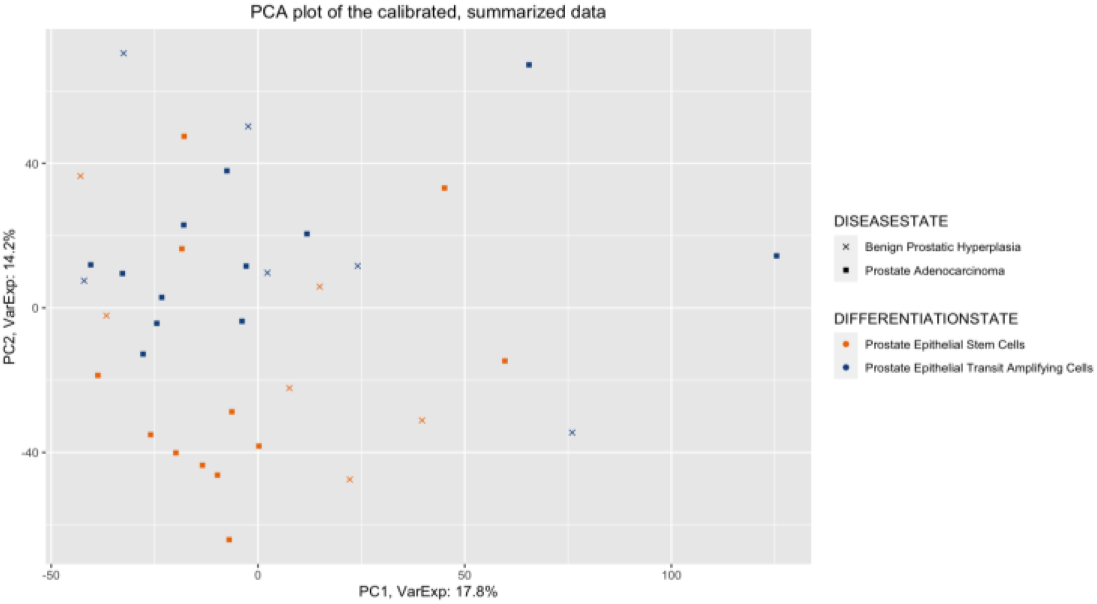
PCA of calibrated data

We now jump into analyzing differentially expressed genes. We use the AnnotationDbi function [19] to query the gene symbols and associated short descriptions for transcript clusters. We filtered out the probes that do not map to a gene. Table 4 shows a sample of Annotated genes.

**Table 4.**
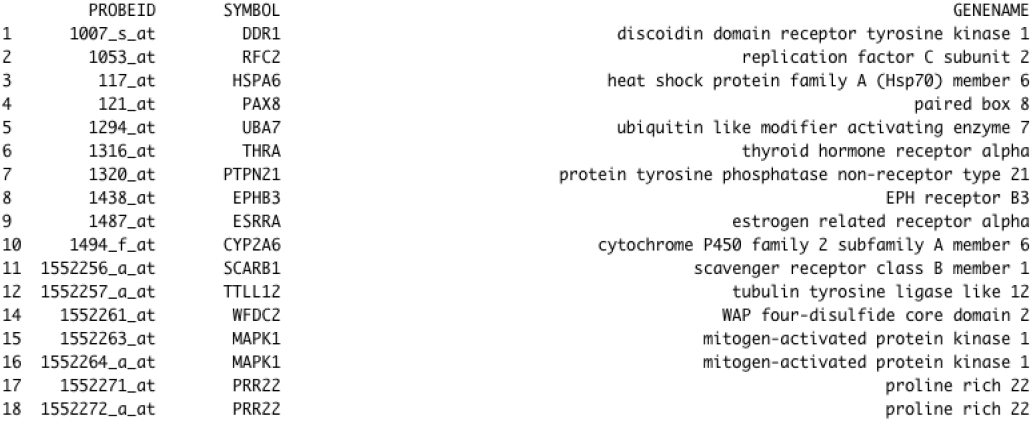
Sample of Annotated Genes

We then create a linear model for our data and check the results for a single gene to check the integrity of the process. We are interested in the changes in the transcription between the two differentiation states.

Before fitting a linear model for the gene expressions of the ADGRF3 gene, we see on the top of figure 9 that the gene expression between the various differentiation states for patients with prostate adenocarcinoma is not clear. After fitting our linear model, we can plot the expression values again, and we notice that for patients with PA, Prostate epithelial transit amplifying cells is expressed slightly higher on the bottom of figure 8.

**Figure 9.**
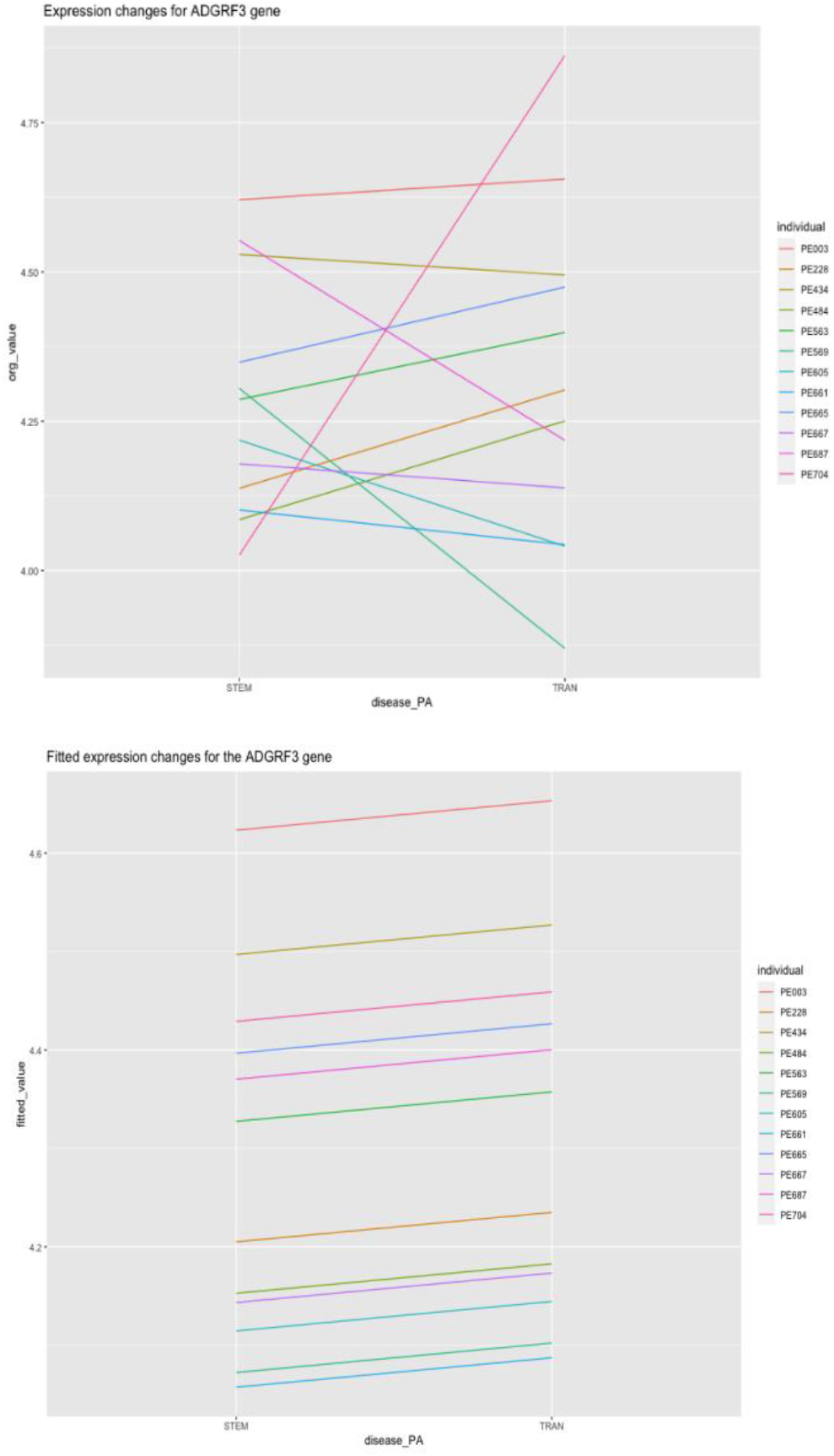
ADGRF3 expression change vs Fitted Expression change

However, In order to further test if gene ADGRF3 is differentially expressed or not we need to run a t-test with the null hypothesis that there is no difference in the expression between the two differentiation states. We get a high P-value (0.58) which indicated this gene is not differentially expresses between the two differentiation states. Table 5 shows the full output of our t-test on gene ADGRF3.

**Table 5.**
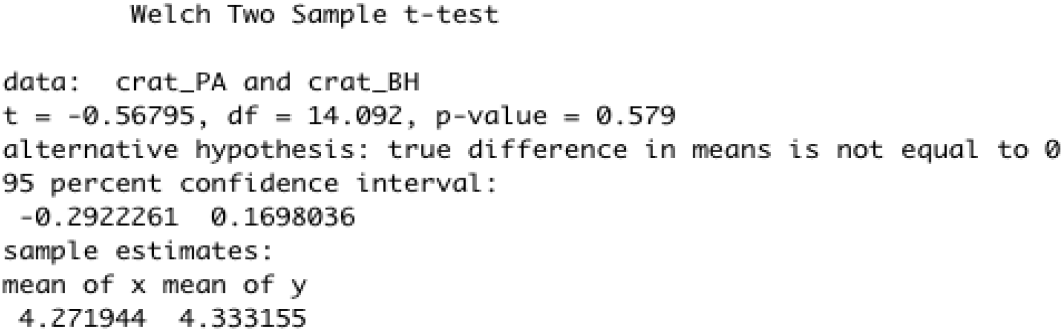
Two Sample T-test for ADGRF3 gene

Next step is for us to fit our linear model to all the genes and define appropriate contrasts to test hypotheses of interest. We apply the empirical Bayes variance moderation method to the model that computes moderated t-statistics. We showcase the results of P-values for prostate adenocarcinoma with the two differentiation states in a histogram in figure 10. Peak near zero shows enrichment for low p-values corresponding to differentially expressed genes. The total number of differentially expressed genes for PA is 1347, and the total number of differentially expressed genes for BPH is 71.

**Figure 10.**
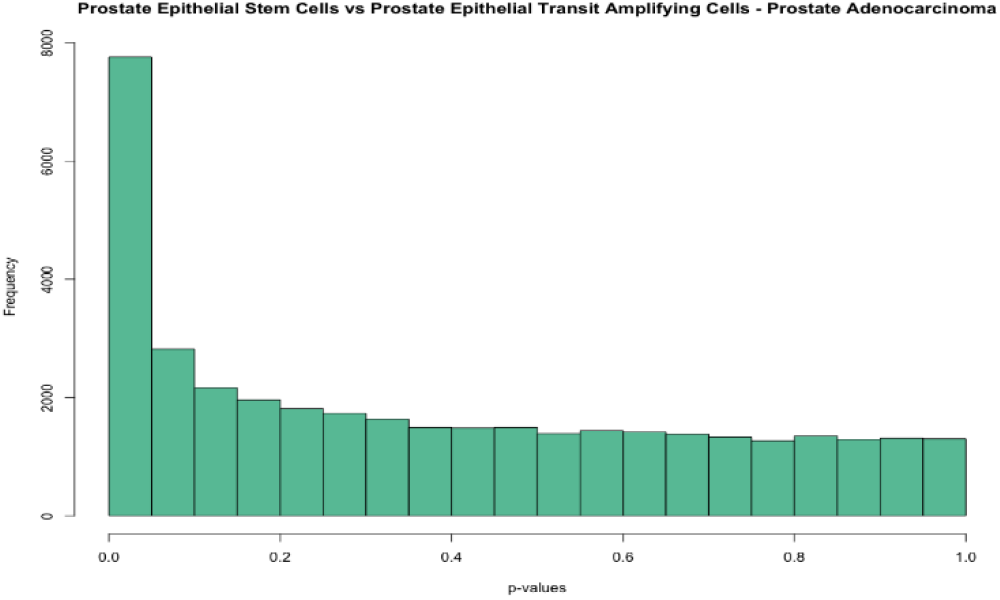
p-value frequencies

In order to visualize differentially expressed genes, we draw a volcano plot, figure 11. We only show gene names with a fold change greater than 1.

**Figure 11.**
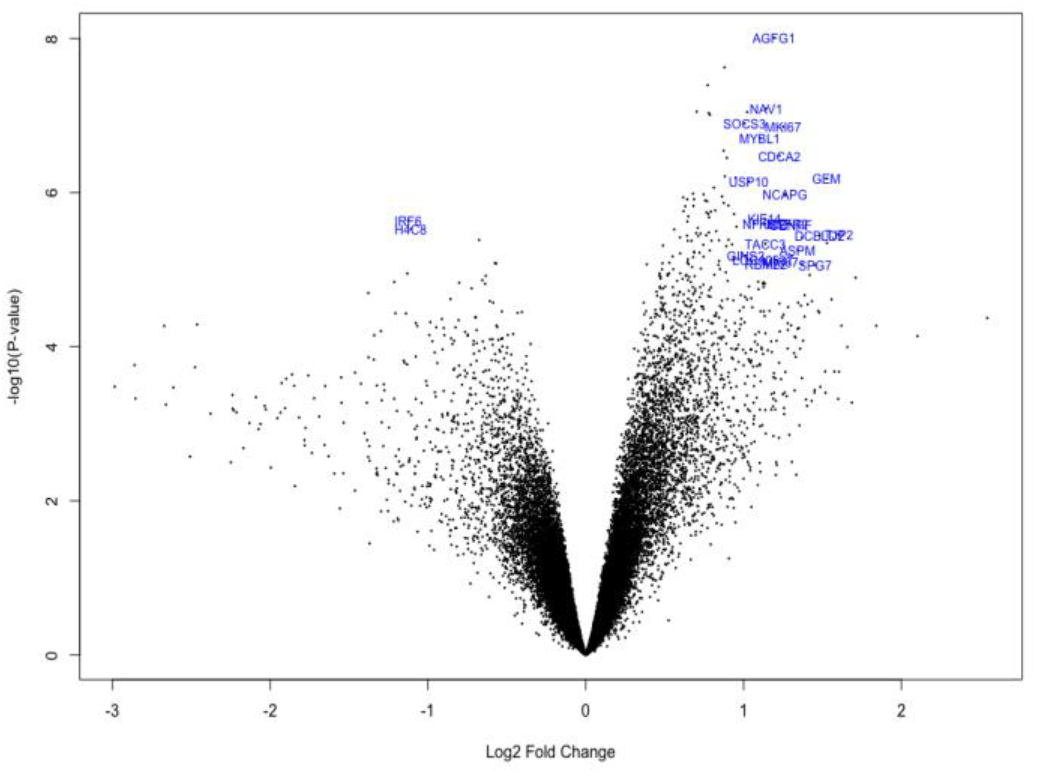
Volcano Plot of Genes

The USP10 gene encodes a protein called ubiquitin carboxyl-terminal hydrolase 10, which plays a role in regulating the stability of other proteins in cells [20]. Some studies have suggested that USP10 may be involved in the development of prostate adenocarcinoma, the most common type of prostate cancer [21].

The SOCS3 gene encodes a protein called suppressor of cytokine signaling 3, which plays a role in regulating the immune system [22]. Some studies have suggested that the SOCS3 gene may be involved in the development of prostate adenocarcinoma [23].

## IV. Conclusion

The total number of differentially expressed genes for patients battling prostate adenocarcinoma is 1347. The total number of differentially expressed genes for patients battling benign prostatic hyperplasia is 71.Genes USP10 and SOCS3 were found to be differentially expressed and are found to be associated with prostate cancer.

## Data Availability

All data produced are available online at https://www.ebi.ac.uk/biostudies/arrayexpress/studies/E-MEXP-993?query=E-MEXP-993#].

## Notes

### Competing Interest Statement

The authors have declared no competing interest.

### Funding Statement

This study did not receive any funding

